# A Rest-Shade-Hydration-Hygiene program reduces acute kidney injury and increases production at a sugar mill in Nicaragua, an economic analysis

**DOI:** 10.1101/2025.02.19.25322486

**Authors:** Zachary J. Schlader, Thomas Boswell, Heath Prince, Catarina Wesseling, Fabiano A. Amorim, Dinesh Neupane, Esteban Arias, Scarlette Poveda, Erik Hansson, Rebekah A.I. Lucas, Kristina Jakobsson, David H. Wegman, Jason Glaser

## Abstract

**Background:** Occupational heat stress mediated acute kidney injury (AKI) has been linked to the development of chronic kidney disease of non-traditional causes (CKDnt) in agriculture workers. Rest-shade-hydration-hygiene (RSHH) programs are promising interventions for preventing CKDnt. An obstacle to the implementation of RSHH programs is the concern that the reduced work time associated with these programs may reduce productivity and earnings. This case study analyzes the economic impact of an RSHH program implemented at a sugar mill in Nicaragua.

**Approach:** Data were obtained from the sugar mill over a six-year, five-harvest period (Harvest 1: 2017-18 through Harvest 5: 2021-2022). Data included health and productivity metrics, and RSHH program costs. During Harvest 1 existing heat mitigation strategies were in place but were not optimal. Thus, 2017 was considered the pre-RSHH (baseline) period. Over subsequent harvests progressively improved RSHH programs were implemented.

**Analysis:** A cost-benefit analysis was conducted to estimate the return on investment of the RSHH program. The analysis considered both fixed and variable costs associated with the program, including electrolyte beverage production, costs for AKI treatment and worker training. Benefits were calculated based on productivity improvements, including reductions in absenteeism, and reductions in AKI cases.

**Results:** As soon as 2020, the costs of implementing the RSHH program were offset by savings resulting from increased productivity (i.e., reduced absenteeism and increased worker production) and reduced cases of AKI. The RSHH program yielded a positive return on investment from 2020 and onward. The average return on investment over the five-year period was 0.02 (or a return of $1.02 for every $1.00 invested), which takes into consideration the first two years of the intervention in which there was a negative return on investment. In 2022, every $1.00 invested in the RSHH program saw a return of $1.60.

**Discussion:** This case study provides evidence that implementing an RSHH program can provide both economic and health benefits, particularly in locations where climate change is increasing the already present risk and burden of occupational heat stress.

**Teaser message:** A rest-shade-hydration-hygiene intervention program at a sugar mill in Nicaragua produced a positive return on investment after five years by improving both health and productivity outcomes.

**Key findings and implications:**

- A positive average return on investment of $1.60 USD for every $1.00 invested was identified after a five-year period following implementation of a rest-shade-hydration-hygiene (RSHH) program at a sugar mill in Nicaragua.
- Three years after implementation, the costs of implementing the RSHH program were offset by savings resulting from increased productivity (i.e., reduced absenteeism and increased worker production) and reduced cases of acute kidney injury.
- This case study provides evidence of the health and economic benefits of promoting an easily applicable workplace intervention that addresses current risks as well as the rising threat of climate change induced occupational heat stress.

## Background

The most evident manifestation of climate change is the heightened intensity, frequency, and duration of heat exposure ^1^. Less well recognized are the risks posed by this heat exposure when combined with the physical activity required by many occupational tasks, which renders outdoor workers uniquely and disproportionately vulnerable to the health impacts of climate change ^2^. Seemingly occurring in parallel with climate change, epidemics of chronic kidney disease not related to traditional causes of diabetes, hypertension, obesity, or specific kidney diseases (CKDnt) have emerged globally, including in Mesoamerica ^3,4^. A leading hypothesis is that occupational heat stress, a function of both environmental heat exposure and intense manual labor ^5^, catalyzes the development of CKDnt following single or repeated episodes of heat-induced acute kidney injury (AKI) ^6^. Thus, CKDnt is arguably the first occupational disease recognized to be caused by climate change ^7^. This occupational heat stress mediated AKI-to-CKDnt hypothesis contends that elevations in internal (core) body temperature increase the risk of developing AKI, which is worsened in the presence of dehydration ^5^. This hypothesis is supported by: (i) a high CKD mortality in areas where excessive occupational heat stress is common ^8^; (ii) clinical data demonstrating that AKI caused by heat stroke increases the likelihood of developing CKD ^9^; (iii) data from rodents demonstrating that CKD can be caused by repetitive bouts of heat-induced AKI ^10,11^; (iv) positive associations between AKI and the diagnosis of CKDnt in sugarcane workers ^12^, and (v) the introduction of a rest-shade-hydration-hygiene (RSHH) program that reduces dangerously high core temperatures, reduces the incidence of AKI, and attenuates the fall in kidney function across the harvest in sugarcane cutters ^13–15^. To this latter point, despite data supporting the beneficial renal health effects of an RSHH program, a major obstacle for RSHH program success is that both employers and workers often worry that reduced work time with the RSHH program will decrease productivity and earnings.

It is well established that occupational heat stress is a major economic burden. For example, occupational heat stress accounts for an estimated >650 billion lost labor hours annually worldwide, comparable to those caused by the COVID-19 pandemic ^16^, and this burden is expected to worsen with climate change ^17^. However, estimates of the financial benefits of implementing RSHH programs to counter the impacts of occupational heat stress are preliminary to date. Indeed, initial estimates indicate that worker productivity (e.g., tons of sugarcane cut) decreases with increased heat stress but that implementation of RSHH reverses these decrements, such that despite reductions in functional working time productivity is maintained ^18^. Thus, RSHH programs likely have positive economic consequences. That said, aside from preliminary analyses conducted by our group ^19,20^, a comprehensive economic assessment of the potential benefits of an RSHH program incorporating adverse health outcomes (specifically AKI), productivity, and RSHH program fixed and annual costs has not been undertaken.

This case study analyzes the economic impact of an RSHH program, the Adelante Initiative, implemented at the Ingenio San Antonio (ISA) sugar mill in Chichigalpa, Nicaragua (https://adelanteinitiative.org/). Chichigalpa is in a region with one of the highest known rates of CKDnt in the world ^8^. The weather in Chichigalpa is hot and humid ^8^, and the frequency and severity of extreme heat in the region is predicted to increase due to climate change. ISA is a large sugar mill with complex production processes, including both manual and mechanized harvesting. This case study focuses on manual harvesting, which, despite mechanization, is and will remain a requirement at ISA due to the terrain, sugarcane, and other production conditions.

## Approach

The human subjects aspects of this case study were approved by the Comité de Ética para Investigaciones Biomédicas (CEIB), Facultad de Ciencias Médicas, Universidad Nacional Autónoma de Nicaragua (UNAN-León), FWA000045231/IRB00003342. In all instances, trained staff apprised all workers of the study objectives and procedures and answered any questions before participants signed an informed consent.

This case study presents an economic assessment of the RSHH program implemented for cane cutters (both those cutting burned cane and those cutting cane for seed (seed cutters) at ISA as assessed over five harvest periods, with data presented from 2017-2018 (Harvest 1) through 2021-2022 (Harvest 5). Data included health and productivity metrics, and RSHH program fixed and annual costs (described in detail below). The initial observations, development, and implementation of the RSHH program at ISA have been described previously in detail ^19,21^. Briefly, existing heat mitigation strategies were examined among sugarcane workers during Harvest 1 of the Adelante Initiative. During the harvest, existing heat mitigation strategies were in place but were not considered to be ideal. For example, water and shade were available but these resources were not convenient for workers to access ^19^. Thus, for the purposes of this case study, 2017-2018 is considered the pre-RSHH (baseline) period (Harvest 1). Based on the findings in Harvest 1, an improved intervention was designed and deployed during Harvest 2 (2018-2019) ^19,21^. The enhanced RSHH program during Harvest 2 included the following improvements: (i) increased rest time - for burned cane cutters, a regulated rest schedule was implemented during a six-hour workday, with an additional 20 minutes of rest more evenly distributed throughout the day. For seed cutters, the workday remained eight hours but with two additional rest periods; (ii) improved access to shade and fluids - large shade tents and palatable fluids were placed close to workers (within 50 meters) throughout the day; (iii) delayed cutting after burning - cutting was delayed for at least 12 hours after a field was burned to reduce exposure to radiant heat and other pollutants; (iv) hygiene (bathroom) facilities were placed in the field, which is particularly necessary for women who otherwise tend to restrict their liquid intake to avoid urination, thereby increasing their risk for dehydration and heat-illness. The RSHH program was further modified in Harvest 3 (2019-2020) to add breaks earlier in the day for the burned cane cutters and increase break periods for seed cutters. Assessment and improving organization management to optimize the implementation of the RSHH program also became a focus in Harvest 3 ^19,21^. The COVID-19 pandemic occurred during Harvest 3, but sugarcane harvesting continued. The workers were required to wear facemasks, were specifically instructed to wash their hands before and after work shifts, were requested to keep physical distance, and were monitored for signs and symptoms of COVID-19, with testing provided by mill hospital staff as needed. Aside from modest improvements in the implementation, the RSHH program was largely unchanged in Harvest 4 (2020-2021) and Harvest 5 (2021-2022).

ISA provided the research team with implementation and operational costs and worker outcomes data. These are data that ISA regularly collects to monitor production, employee turnover, absenteeism, and harvest-working days. ISA was blind to the purpose of providing these data to the research team. While the RSHH intervention was designed to target those workers at highest risk of occupational heat stress (e.g., burned cane cutters, seed cutters), the obtained data specifically included information related to the annual total costs of implementing the RSHH program at ISA more broadly and were not specific to a given worker group. Costs included were specifically related to the production and distribution of an electrolyte solution (including ingredients and packaging, salaries for personnel, electricity consumption), equipment for new workers (e.g., hats, thermoses, shade tents, water reserves, etc.), worker training on occupational heat stress health risks and its prevention (i.e., a cost per worker) and salaries for personnel responsible for the implementation of the RSHH intervention (e.g., health promotion employees). Fixed costs that were provided to the research team only one time from 2017 through 2022 were adjusted for inflation to estimate annual fixed costs. The research team was also provided with the total number of workers (including the number of burned cane cutters), burned cane cutter productivity measures (i.e., tons of sugarcane cut), absenteeism, and the number of AKI cases in burned cane cutters, including the cost for treatment at the mill’s on-site hospital. From the variable cost data, we calculated the cost savings due to the RSHH program using data from 2017 (pre-RSHH) as baseline. Cost savings data focused specifically on burned cane cutters given the ability to directly align burned cane cutter productivity with a monetary value (more information provided below). The use of ISA-wide costs relative to burned cane cutter specific cost savings is an acknowledged limitation. However, this was deemed acceptable because we will be underestimating (and not overestimating) the cost savings. A summary of the data provided and used in the cost-benefit analysis has been included in **Table 1**. All amounts are presented in U.S. dollars.

**Table 1:** Variables employed in the cost-benefit and return on investment calculations.

| **Fixed RSHH program costs (ISA wide)** |  |
| --- | --- |
| Electrolyte beverage production |  |
| Salaries for intervention staff |  |
| Worker employment and training costs |  |
| Worker equipment costs |  |
| **Variable RSHH program outcomes costs \*** | **Cost savings due to RSHH program \*** |
| Cost of AKI treatment | Reduced need for AKI treatment |
| Cost of absenteeism (i.e., worker turnover) | Reduced absenteeism |
| Cost of productivity loss | Increased daily sugarcane cut |

\* Data specific to burned cane cutters only.

## Analyses

### Value of investments

#### Electrolyte Beverage Production

ISA provided a detailed cost estimate for their operations related to electrolyte beverage production and distribution. These costs can be broken down into three categories: ingredients and packaging ($140,462.36), operation salaries ($53,788.09), and energy usage (electricity) to produce and maintain the beverage ($32,294.43), which totaled $226,544.88 in 2020. These costs were subsequently adjusted for inflation to determine annual fixed costs in 2017 through 2022.

#### Equipment Costs

Prior to 2017, subcontracting during the harvest period was regularly used at ISA, as in many other sugar mills in the region. In 2017 ISA eliminated subcontracting. With formalization of the workforce and establishment of an employer-employee working relationship, ISA invested in providing uniform equipment of specified quality for its cutters, some of which was important in implementing the RSHH program. This equipment included making annual purchases of hats, shirts, shin guards, gloves, machetes, thermoses, water reserves, and shade structures. While there is no need to purchase new equipment for employees each harvest season, ISA estimates that these annual purchases could equip 180 workers per harvest to account for equipping new employees and replacing degraded equipment. ISA provided estimates on the total value of these purchases by category in 2022, amounting to $34,438.00. These costs were subsequently adjusted for inflation to determine annual fixed costs in 2017 through 2021.

#### Annual Occupational Safety and Health Training

The Occupational Health unit at ISA conducted training for the priority jobs in the RSHH intervention (burned cane cutters, seed cutters) to review practices and ensure workers (and their supervisors) were adopting the RSHH program appropriately to reduce the incidence of heat-strain and associated illness ^19^. The value of this training includes the dollar value of time dedicated per-harvest cycle and the materials produced to educate and refresh workers. In 2022, we estimate the cost per worker to be $7.21 at a total value of $2,617.23 (363 total burned cane and seed cutters in 2022). These costs were subsequently adjusted for inflation to determine annual fixed costs in 2017 through 2021.

#### Health Promotion Staffing

A substantial portion of the intervention costs are attributed to ISA’s mill hospital employees dedicated to the promotion of RSHH-related activities. We include the salaries for three health promotion officials totaling $51,308.65 in 2022. These costs were subsequently adjusted for inflation to determine annual fixed costs in 2017 through 2021.

### Value of Benefits

#### Productivity

The aim of the RSHH program was to improve occupational conditions for workers with the highest risk of occupational heat stress. After conducting initial rounds of monitoring and analysis to establish recommendations for ISA, the intervention was designed to primarily target two working groups: burned cane cutters and seed-cane cutters. Given the ability to directly align burned cane cutter productivity (i.e., tons of sugarcane cut per worker per year) with a monetary value (see below), the productivity assessments in this case study only include burned cane cutters. We acknowledge that we are likely underestimating the potential productivity benefits of the RSHH program as, for example, our group has recently also identified improvements in seed cutters ^18^, not to mention other working groups where the quantification of productivity and subsequent financial implications is less straightforward. Although a limitation, we believe this is acceptable given that only including burned cane cutter productivity is an underestimate of the potential productivity benefits.

Using the data provided by ISA’s human resources department, we can estimate the total number of working days between 2017 and 2022 by multiplying the average number of workers per year by the total number of working days for burned cane cutters. **Table 2** provides a breakdown of average number of workers and total working days between 2017 and 2022 for the burned cane cutters and all employees.

**Table 2:** Average number of burned cane cutters and total number of workers at ISA, and total working days for burned cane cutters.

| Year | Total number of burned cane cutters | Total labor force at ISA | Total number of working days per burned cane cutter |
| --- | --- | --- | --- |
| 2017 | 498 | 3775 | 143 |
| 2018 | 227 | 3503 | 136 |
| 2019 | 180 | 2755 | 124 |
| 2020 | 180 | 2527 | 101 |
| 2021 | 169 | 3028 | 123 |
| 2022 | 154 | 2932 | 156 |

Using findings from our previous study conducted to estimate changes in production for burned cane cutters after RSHH program implementation ^18^, measures of total potential production can be estimated in terms of cane-tonnage harvested. Using this information, average yearly productivity can be calculated from the number of workers and the total number of working days for burned cane cutters. These data are presented in **Table 3**.

**Table 3:** Daily productivity for burned cane cutters from 2017-2022.

| Year | Average sugarcane cut per worker per day (in tons) |
| --- | --- |
| 2017 | 5.7 |
| 2018 | 5.7 |
| 2019 | 5.6 |
| 2020 | 6.3 |
| 2021 | 5.9 |
| 2022 | 6.2 |

To estimate the value of savings from increased production, we used absenteeism rates for burned cane cutters to calculate lost production based on potential productivity. ISA monitors the number of absenteeism days for workers across worker categories. Reasons for missing work are recorded in the following categories: unexcused absences, common illness, unpaid leave, and occupational risk. Table 4 provides a breakdown of the number of recorded working days absent by type of absence for the burned cane cutting crew. While the RSHH program primarily focused on outcomes most closely associated with occupational heat stress, it is likely that the intervention improved working conditions more generally, which had impact on other absenteeism categories such as unexcused absences and common illness. The logic behind this assumption is that if working conditions improve, workers will be more willing to maintain a regular working schedule in the presence of a safe and more welcoming working environment. Using the absenteeism figures provided by ISA, we calculated the total production loss due to missing workdays as the quotient of the total potential sugarcane harvest and lost production (**Figure 1**).

**Figure 1:**
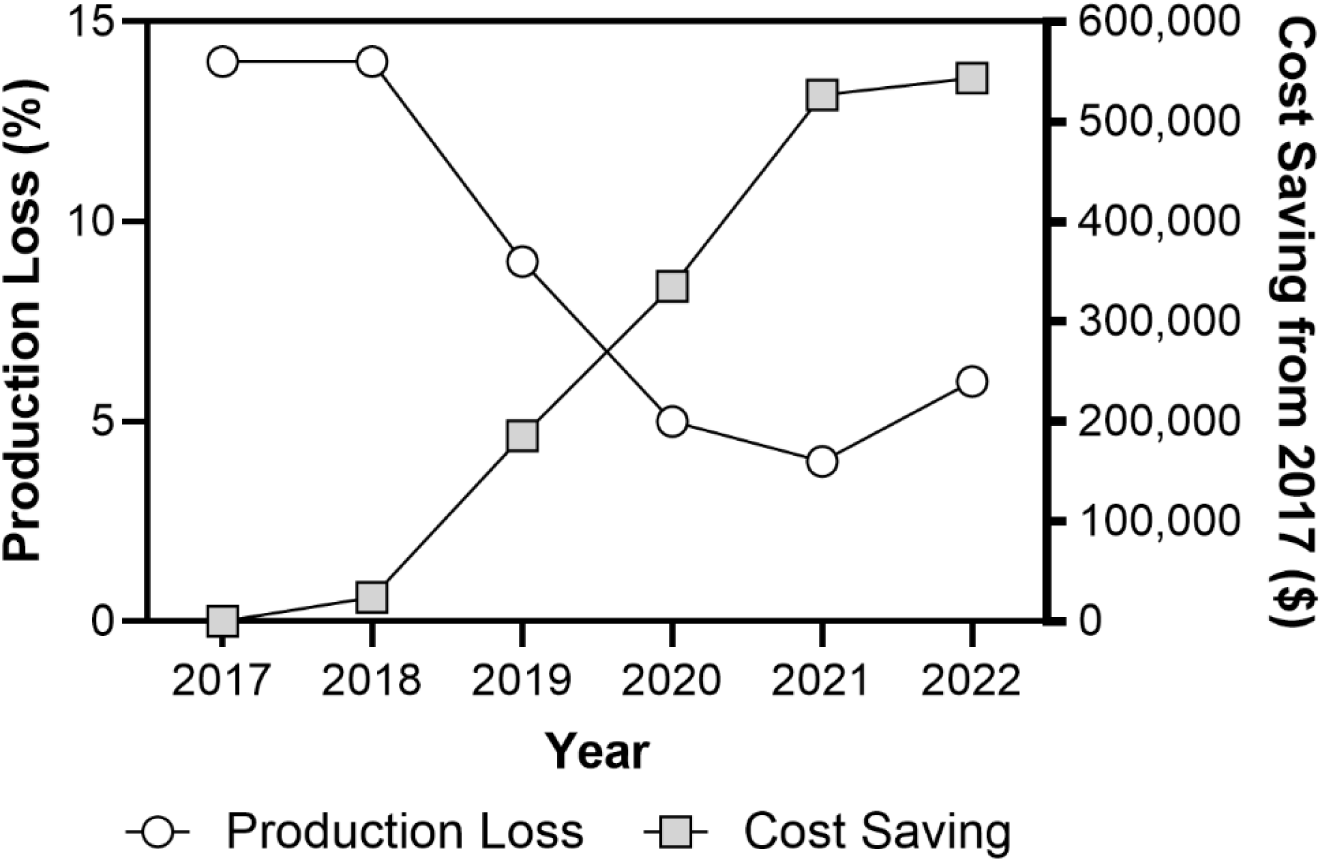
Calculated production losses and cost savings due to enhanced productivity from 2017-2022 in burned cane cutters.

**Table 4:**
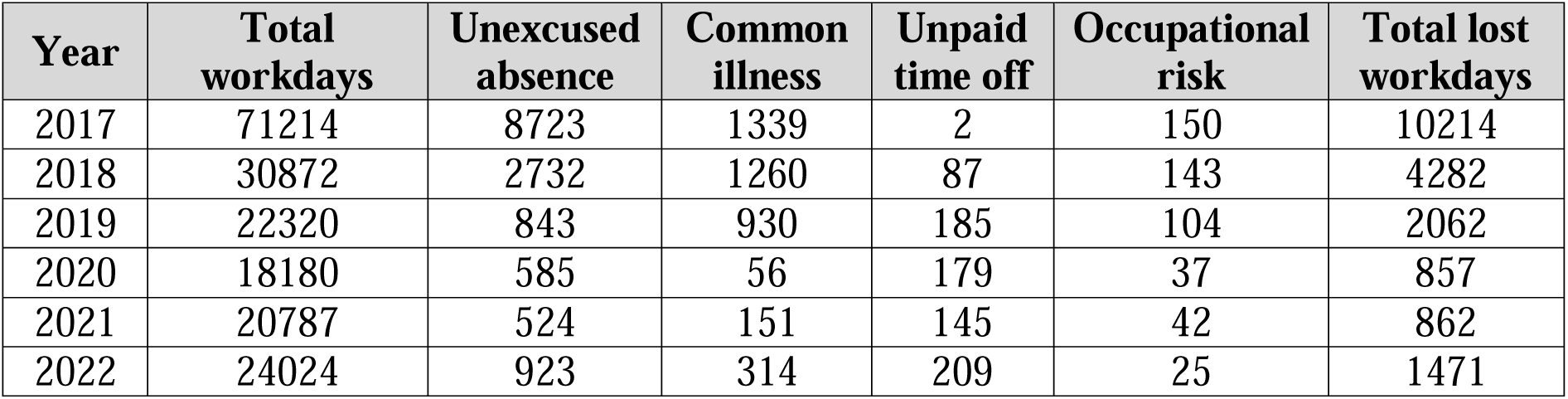
Total workdays, missed workdays by reason, and total lost workdays for burned cane cutters.

| Year | Total workdays | Unexcused absence | Common illness | Unpaid time off | Occupational risk | Total lost workdays |
| --- | --- | --- | --- | --- | --- | --- |
| 2017 | 71214 | 8723 | 1339 | 2 | 150 | 10214 |
| 2018 | 30872 | 2732 | 1260 | 87 | 143 | 4282 |
| 2019 | 22320 | 843 | 930 | 185 | 104 | 2062 |
| 2020 | 18180 | 585 | 56 | 179 | 37 | 857 |
| 2021 | 20787 | 524 | 151 | 145 | 42 | 862 |
| 2022 | 24024 | 923 | 314 | 209 | 25 | 1471 |

It is conservatively estimated that one ton of harvested sugarcane produces ∼107 kg of raw sugar ^22^. The average world market price for 1 kg of processed sugar in each year was determined as reported from the International Monetary Fund’s global price index ^23^. From this information, the monetary value of improvements in total production losses (termed cost savings) can be calculated for each year relative to 2017, with the market price adjusted for inflation (**Figure 1**).

#### Health outcomes

The primary adverse health outcome assessed was AKI. This assumes that AKI in this setting can be considered a heat-related illness ^2^. Moreover, AKI is hypothesized to be central to the development of CKDnt ^6^. The total number of AKI diagnoses in burned cane cutters decreased over time (**Figure 2**), from occurring in one out of every five workers in 2017 to one out of every 10 workers in 2022, as has been reported previously ^15^. Thus, reductions in the occurrence of AKI can be expressed as savings to ISA. According to data provided by ISA in 2020 the total treatment cost for a worker diagnosed with AKI at the mill hospital was $253.48 and this value was adjusted for inflation for years 2017-2019 and 2021-2022. To capture savings associated with a reduction in costs for treating AKI, we took the per-capita burned cane cutter AKI incidence amongst the working population at ISA in all years from 2017 to 2022. Cost savings was calculated as a function of the difference between treatment costs each year using the observed rate and what the observed rate might have been in the absence of the intervention (i.e., 2017) with the AKI treatment cost adjusted for inflation (**Figure 2**).

**Figure 2:**
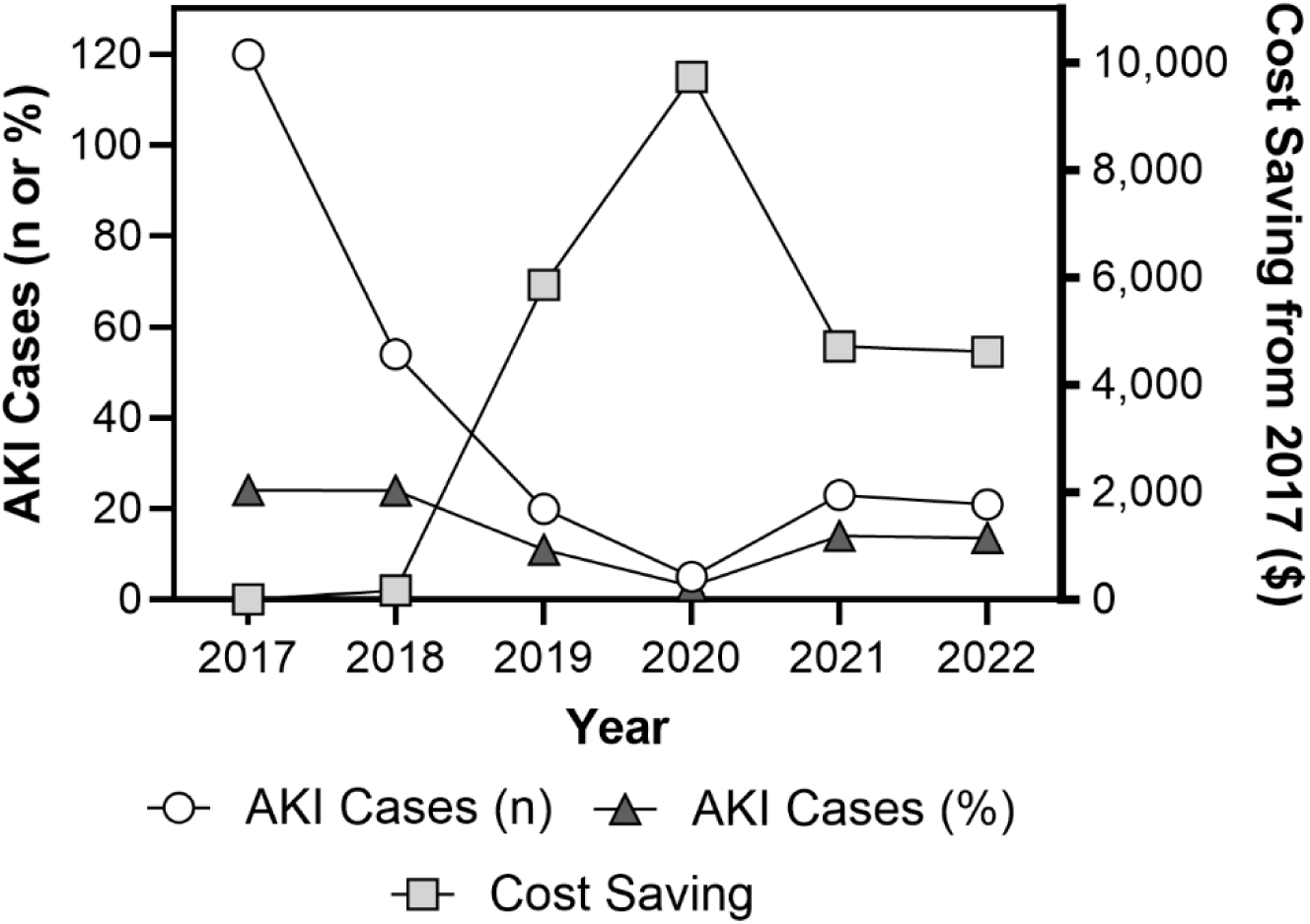
Total number of acute kidney injury (AKI) cases in burned cane cutters (n) and as a function of the number of burned cane cutters (%), and the associated cost savings due to reduced AKI incidence in burned cane cutters.

### Return on Investment (ROI)

The dollar value of expected cost reductions was expressed as firm-level benefits that were then used to conduct a return on investment assessment using the following standard ROI formula ^24^.

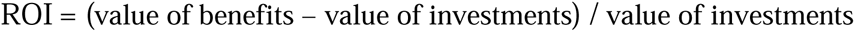

Where the value of investments is the sum of the fixed costs including those costs related to equipment, training, operations, and electrolyte beverage production and the value of benefits is the sum of the cost savings from improvements in productivity and reductions in the incidence of AKI in burned cane cutters.

Given that the value of the benefits was calculated relative to 2017 (i.e., pre-RSHH program implementation), an ROI could not be calculated for 2017. Therefore, ROI is presented as relative to that occurring in 2017.

### Findings

#### Value of investments

Annual fixed costs for equipment, training, operations, and electrolyte beverage production are presented in **Table 5**.

**Table 5:** Annual fixed costs for implementation of the rest-shade-hydration-hygiene intervention.

| Year | Equipment | Training | Operations | Electrolyte beverage production | Total Costs |
| --- | --- | --- | --- | --- | --- |
| 2017 | \$28,927.92 | \$4,530.19 | \$43,098.72 | \$215,217.23 | \$291,774.05 |
| 2018 | \$29,616.68 | \$2,957.69 | \$44,124.88 | \$219,748.12 | \$296,447.36 |
| 2019 | \$29,961.06 | \$2,302.08 | \$44,637.96 | \$224,279.01 | \$301,180.11 |
| 2020 | \$30,305.44 | \$2,525.23 | \$45,151.04 | \$226,544.45 | \$304,526.16 |
| 2021 | \$32,027.34 | \$2,487.67 | \$47,716.44 | \$237,871.67 | \$320,103.12 |
| 2022 | \$34,438.00 | \$2,617.23 | \$51,308.00 | \$255,995.23 | \$344,358.46 |

#### Value of benefits

Annual cost savings due to changes in production (i.e., improvements in productivity) and the treatment of AKI are presented in **Table 6**.

**Table 6:**
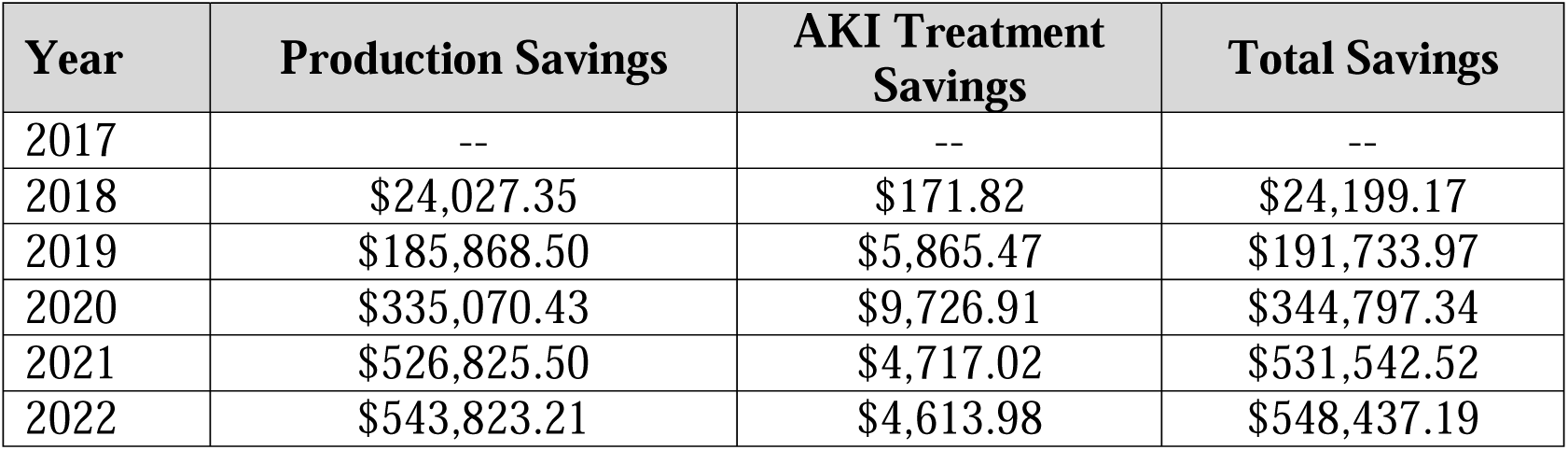
Annual cost savings following implementation of the rest-shade-hydration-hygiene intervention in 2017.

| Year | Production Savings | AKI Treatment Savings | Total Savings |
| --- | --- | --- | --- |
| 2017 | -- | -- | -- |
| 2018 | \$24,027.35 | \$171.82 | \$24,199.17 |
| 2019 | \$185,868.50 | \$5,865.47 | \$191,733.97 |
| 2020 | \$335,070.43 | \$9,726.91 | \$344,797.34 |
| 2021 | \$526,825.50 | \$4,717.02 | \$531,542.52 |
| 2022 | \$543,823.21 | \$4,613.98 | \$548,437.19 |

#### Return on Investment

The annual value of benefits, value of investments and return on investment for the implementation of the RSHH program are presented in **Figure 3**. From 2020 there was a positive return on investment, which escalated with improvements in RSHH intervention implementation.

**Figure 3:**
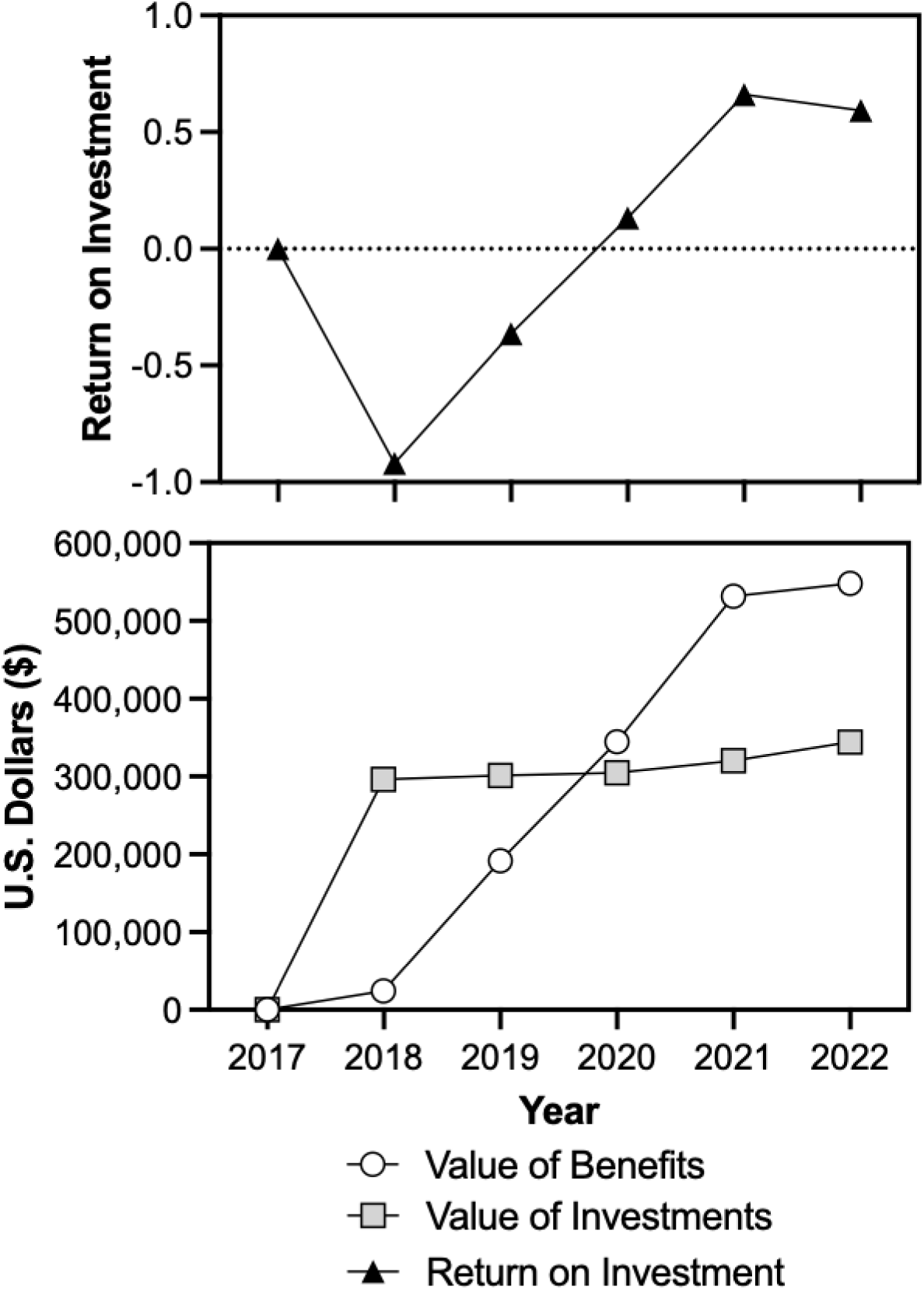
The value of the benefits and investments following the implementation of the rest-shade hydration-hygiene program in 2018 (bottom) and quantification of the annual net return on investment (top).

## Discussion

The purpose of this case study was to determine the economic impact of an RSHH program implemented at the ISA sugar mill in Chichigalpa, Nicaragua. Chichigalpa has one of the highest known rates of CKDnt ^8^, which has been proposed to be predominantly caused by occupational heat stress induced AKI ^6,25^. Implementation of the RSHH program at ISA has already been shown to improve worker productivity ^18^, reduce the incidence of AKI ^15^, and attenuate cross harvest reductions in kidney function ^13,14^. While preliminary evidence supports a beneficial economic impact of the implementation of an RSHH program ^19,20^, a comprehensive economic assessment of the potential financial benefits of an RSHH program were previously lacking. To this end, our analysis demonstrates that as soon as three years following implementation of the RSHH program the health and productivity benefits exceed the costs such that, for example, in 2022 for every $1.00 invested in the RSHH program ISA saw a return of $1.60 (or a net return of $0.60) (**Figure 3**). Notably, even the average return on investment over the five-year period was positive (0.02 or a return of $1.02 for every $1.00 invested), which takes into consideration the first two years of the intervention in which there was a negative return on investment.

Given the vulnerability of outdoor workers to severe heat, which is being made worse by climate change, there is widespread acknowledgement of the need to provide RSHH to outdoor workers to mitigate the short- (e.g., heat illness, AKI) and longer- (e.g., CKDnt) term health and safety implication of occupational heat stress ^2^. However, a major obstacle for successful integration of RSHH programs into the workplace is that both employers and workers worry that the reduced work time in the RSHH program (i.e., increased rest breaks) will reduce workplace productivity and employee earnings, particularly when the work is piece-paid. Overall, the results presented in this case study suggest that daily lost functional work time associated with RSHH for the workers is offset by savings in (i) productivity, due to both reductions in absenteeism in increases in annual per worker harvested tonnage, and (ii) AKI treatment costs due to reductions in the incidence of AKI (**Figure 2**). Notably, despite implementation of the RSHH program, which introduced additional rest resulting in a somewhat shorter overall time working, we observed modest improvements in burned cane cutter productivity (i.e., tons of sugarcane cut, **Table 3**, ^18^). Given that burned cane cutters are piece-paid workers, this suggests that earnings are (at worst) not changed and that the workers are healthier (**Figure 2**). Thus, these findings indicate that there are financial and health benefits to both the employer and employee due to the implementation of the RSHH program.

It is worth noting that the observed economic benefits of the RSHH program are likely an underestimate given the sole focus on burned cane cutters in the determination of cost savings. This was chosen due to the relative simplicity linking burned cane cutter productivity (i.e., tons of sugarcane cut per worker per year) with a monetary value, the latter of which is a function of the yield of sugar per ton of sugarcane and the market price of sugar. We note that other groups of workers likely also displayed improvements in productivity. For example, Hansson et al. ^18^ recently reported an average of an ∼11% increase in productivity in seed-cutters following implementation of the RSHH program at this sugar mill. It is entirely possible that other working groups at ISA also experienced improvements in productivity. Notably, quantification of worker productivity is a challenge that is not exclusive to the sugarcane industry. Thus, future efforts should aim to improve the assessment and reporting of jobsite and worker productivity to better improve economic analyses before and after implementation of RSHH programs.

Aside from initial preliminary assessments ^19,20^, to our knowledge a similar comprehensive economic assessment of the implementation of an RSHH program has not been carried out. Thus, it is reasonable to consider whether the observed economic returns following the implementation of the RSHH program is specific to ISA, a large sugar mill in Nicaragua, a lower middle-income country in Central America. It is well established that occupational heat stress reduces worker productivity ^26^ and increases the risk of heat illness, including AKI ^27^. Thus, the observations made in this case study are likely transferable to other industries, job types and locations where occupational heat stress is common. Nevertheless, it is impossible to ignore the potential that the economic effects of an RSHH program may not be as profound in other climate regions where occupational heat stress is more seasonal (e.g., at higher or lower latitudes), for employers in more economically prosperous regions and in different industries, particularly those not heavily relying upon piece-paid work. Therefore, future research should examine the potential effects of RSHH programs on both employee and employer economic returns in more diverse settings, as this information would be vital towards widespread adoption of RSHH programs.

The focus of the current case study has been on the short-term economic implications of the implementation of an RSHH program. That said, there may also be longer term societal benefits. For example, given the links between occupational heat stress, AKI and CKDnt ^6^ it is likely that a proportion of those experiencing AKI will develop CKDnt. With its outsized impact on working-age men, CKDnt contributes to lost economic opportunity for households and communities ^28^. There are also benefits to state-funded entities like the Nicaraguan social security and public health systems that provide pensions to sick workers. By reducing the incidence of AKI and CKDnt, the state can focus its efforts on addressing other public health issues. Therefore, ISA’s implementation of an RSHH program can help mitigate these long-term negative societal effects while still maintaining a productive workforce.

Although clear benefits have been observed, it is important to note the limitations present in this case study. For example, almost all the data used in the analysis come from one source, introducing opportunities for reporting biases that should not be overlooked. Additionally, productivity was estimated using data related to worker totals, working days per activity, and estimates on daily worker production. A more accurate analysis would ideally include financial data provided directly related to tonnage harvested, sugar extraction yields, and revenue from those yields. There are also some other expenditures that were ultimately inaccessible to the research team, particularly related to other relevant health-related outcomes (e.g., injuries, other heat illnesses), which would potentially be impacted by the RSHH program. Future studies should prioritize understanding data limitations and establishing protocols for data collection and sharing to improve understanding of the intervention impact. A final limitation is that this case study simply compares the before- and after-effects of the RSHH program. Thus, more sophisticated designs (e.g., those employing randomization, addressing certain confounders, including control groups, etc.) are required to better infer that the RSHH program caused the observed economic changes. That said, it should not be ignored that this case study provides a firm foundation from which future work can be carried out.

## Conclusion

The RSHH program at ISA, a sugar mill in Nicaragua, yielded health and productivity benefits that ultimately resulted in large economic returns five years after implementation. The program’s success in reducing AKI and increasing productivity highlights the strategic importance of creating safer working conditions, particularly when under occupational heat stress conditions. The positive return on investment for this intervention after only three years indicates that a RSHH program can likely be applied across the sugarcane industry and perhaps in other industries where occupational heat stress is common, although further research is needed. Future research should also aim to expand this cost-benefit analysis to identify societal returns, and, in the context of publicly funded programs, taxpayer returns for occupational safety and health interventions.

## Data Availability

Data produced in the present study are available upon reasonable request to the authors

## Acknowledgements

This writing project was supported by the National Institutes of Health (NIH) Climate Change and Health Initiative (https://climateandhealth.nih.gov) and coordinated by the Center for Global Health Studies at the Fogarty International Center of NIH. The activity was led by a steering committee of global experts on health and climate change. More information is available at https://go.nih.gov/ClimateAdaptationStudies. We would like to thank all workers from Ingenio San Antonio who are represented in this study, as well as Denis Chavarría and his OSH team who supported our efforts in the field. We also would like to acknowledge and thank the wider La Isla Network team who have supported the RSHH program implementation at ISA.

